# Association of cerebral venous thrombosis with recent COVID-19 vaccination: case-crossover study using ascertainment through neuroimaging in Scotland

**DOI:** 10.1101/2021.08.23.21261779

**Authors:** Paul M McKeigue, Raj Burgul, Jen Bishop, Chris Robertson, Jim McMenamin, Maureen O’Leary, David A McAllister, Helen M Colhoun

## Abstract

**Objectives:** To investigate the association of primary acute cerebral venous thrombosis (CVT) with COVID-19 vaccination through complete ascertainment of all diagnosed CVT in the population of Scotland.

**Design:** Case-crossover study comparing recent (1-14 days after vaccination) with less recent exposure to vaccination among cases of CVT.

**Setting:** National data for Scotland from 1 December 2020, with diagnosed CVT case ascertainment through neuroimaging studies up to 17 May 2021 and diagnostic coding of hospital discharges up to 28 April 2021 and with linkage to vaccination records.

**Main outcome measure:** Primary acute cerebral venous thrombosis

**Results:** Of 50 primary acute CVT cases, 29 were ascertained only from neuroimaging studies, 2 were ascertained only from hospital discharges, and 19 were ascertained from both sources. Of these 50 cases, 14 had received the Astra-Zeneca ChAdOx1 vaccine and 3 the Pfizer BNT162b2 vaccine. The incidence of CVT per million doses in the first 14 days after vaccination was 2.2 (95% credible interval 0.9 to 4.1) for ChAdOx1 and 1 (95% credible interval 0.1 to 2.9) for BNT162b2. The rate ratio for CVT associated with exposure to ChAdOx1 in the first 14 days compared with exposure 15-84 days after vaccination was 3.2 (95% credible interval 1.1 to 9.5). The 95% credible interval for the rate ratio associated with recent versus less recent exposure to BNT162b2 (0.6 to 95.8) was too wide for useful inference.

**Conclusions:** These findings support a causal association between CVT and the AstraZeneca vaccine. The absolute risk of post-vaccination CVT in this population-wide study in Scotland was lower than has been reported for populations in Scandinavia and Germany; the explanation for this is not clear.

**What is already known on this topic:** The risk of cerebral venous thrombosis (CVT) within 28 days of receiving the AstraZeneca ChAdOx1 vaccine has been estimated as 18 to 25 per million doses in Germany and Scandinavia, but only 5 per million doses in the UK based on the Yellow Card reporting scheme. Risk estimates based on adverse event reporting systems are subject to under-ascertainment and other biases.

**What this study adds:** All diagnosed cases of CVT in Scotland were ascertained by searching neuroimaging studies from December 2020 to May 2021 and linked to national vaccination records. The risk of CVT within 28 days of vaccination with ChAdOx1 was estimated as 3.5 per million doses with an upper bound of 6 per million doses, against a background incidence of about 12 per million adults per year. This indicates that the Yellow Card system has not seriously underestimated the risk in the UK; the explanation for higher risk in other European countries is not clear.

## Introduction

The first reports in international media of an association of COVID-19 vaccines with thrombotic events appeared on 7 March 2021, when the Austrian Federal Office for Safety in Health Care announced that it had suspended use of a batch of AstraZeneca ChAdOx1 vaccine after cases of thromboembolic events. On 7 April 2021 the UK Medicines and Healthcare products Regulatory Agency (MHRA) and the Joint Committee on Vaccines and Immunisation (JCVI) concluded that there was “a possible link” between the vaccine and cerebral venous thrombotic events and issued new guidance [1]. By 18 May 2021 two countries in the EU/ EEA had discontinued use of the vaccine and 15 had limited its use to older age groups [2]. In the US cerebral venous thrombosis has also been reported following administration of the AD26.COV2.S Johnson & Johnson (JJ) vaccine which like the AstraZeneca ChAdOx1 vaccine utilises recombinant adenoviral vectors encoding the SARS-CoV-2 spike protein [3]. An underlying syndrome denoted “vaccine-induced immune thrombotic thrombocytopenia” (VITT) [4,5] or “thrombosis with thrombocytopenia syndrome” (TTS) [6] was described, consisting of thrombosis, thrombocytopenia and platelet-activating antibodies to platelet factor 4–polyanion complexes. It was recognized that the spectrum of this syndrome may include thrombocytopenia alone or thrombosis without thrombocytopenia.

Risk / benefit assessments of the use of the AstraZeneca ChAdOx1 vaccine depend on estimating the incidence of CVT and TTS syndrome post-vaccination and evaluating evidence for causality. However, most such estimates have come from voluntary reporting schemes such as the MHRA Yellow Card scheme. These reporting schemes have limitations: vaccine-associated cases may be under-ascertained if the connection with vaccine exposure is not made by the patient or clinician and, once there has been publicity about any possible association with a particular medicine, reporting can be biased thereafter. Although studies based on linking vaccination records to hospital discharge records ascertained either directly[7] or via primary care [[8] are less subject to reporting bias, these estimates of incidence depend on the accuracy of coding of hospital diagnoses. Therefore here we took advantage of a nationwide database that captures radiologists’ reports of all imaging studies conducted in the National Health Service (NHS) in Scotland to obtain an unbiased and comprehensive ascertainment of diagnosed CVT since the start of the vaccination programme, linked to the national vaccination database. The aim of this study was to understand the epidemiology of CVT and its association with COVID-19 vaccination. The specific objectives were:

1. to estimate incidence of diagnosed primary acute CVT in Scotland during the vaccination programme
2. to ascertain whether diagnostic coding of hospital discharges or deaths adequately captures diagnosed CVT and allows differentiation of secondary and chronic cases from primary acute cases.
3. to investigate the association of diagnosed CVT with vaccination.

## Methods

In brief, we retrieved all potentially relevant scan reports from the nationwide Picture Archiving and Communication System (PACS) to identify diagnoses of CVT from the study start date of 1 December 2020 up to 17 May 2021. We also retrieved all hospital discharge and death records with diagnostic codes for CVT in this time period. As all health care records in Scotland use the Community Health Index (CHI) as a unique identifier, we were able to link PACS reports and hospital discharge and death records to the daily updated national vaccination programme database held by Public Health Scotland/ National Services Scotland. We pre-specified a case-crossover design that would compare exposure to vaccine in a recent time window (1 to 14 days since vaccination) with exposure in a less recent time window. The methods are described in detail below.

### Vaccination doses

The COVID-19 vaccination programme in Scotland began on 8 December 2021. Initial priority groups included care home residents and staff, front line health and social care workers and clinically extremely vulnerable individuals. Initially the Pfizer BNT162b2 product (hereafter Pfizer vaccine) was used. From 8 January 2021 the AstraZeneca ChAdOx1 product (hereafter AstraZeneca vaccine) was introduced and from 7 April 2021 the Moderna mRNA-1273 product (hereafter Moderna vaccine) was introduced. CHI number, date of administration, age at vaccination, vaccine product name, whether first or second dose were extracted for all 6894008 vaccination records from 4 December 2020 to 15 July 2021 from the national vaccination database.

### Ascertainment of CVT from CT scan and MRI reports

The key first-line investigation for patients presenting with any acute neurological syndrome is a non-contrast CT scan of the head. However, this is positive in only about 30% of patients with CVT [9]. For this reason current guidance recommends CT or MR venography in all patients where CVT is suspected [10,11] unless the initial CT shows strong evidence of CVT and a venogram is contra-indicated.

Fig S1 shows a flow diagram of the process of neuroimaging report extraction and processing. In the pilot stage of the project 13 April 2021 we initially restricted the query of the PACS database to records where the study type (RIS_CODE field) was coded as CT or MR venogram. Comparison with hospital discharge records indicated that this query was insensitive. After discussion with radiologists the query was therefore broadened to extract reports with a wider range of potentially relevant study type codes as listed in Supplementary Material and any reports with a study modality of CT or MRI where the study description field contained any of the strings “head”, “brain” or “cerebr”. All such reports of CT and MRI studies from 1 Dec 2020 to 17 May 2021 were retrieved from the PACS database.

For three of the 14 health boards in Scotland, the PACS database contained the radiology report only as an image that could not be queried. For these boards extractions were carried out directly from the local radiology information system that feeds the national PACS instead. For Lanarkshire (12.1% of the Scottish population) the same query was run as for the national PACS. From the two other boards that do not encode reports as text in PACS – Forth Valley and Dumfries & Galloway, covering 8.3% of the Scottish population – only the initial RIS code query for venograms up to 13 April 2021 was completed; a subsequent broader query could not be obtained at the time of this study.

The extracted scan reports were filtered to retain any scan likely to be informative for the presence of CVT i.e. all those with a study type or study description including a venogram or where any of the following strings appeared in the the radiologist’s report: “sinus thrombosis”, “sinus thrombus”, “venous thrombosis”, “venous thrombus”, “venogr”. For any individual with at least one scan meeting these criteria all scan reports for that person since 1 Dec 2020 were retained for review. Scan reports include a summary of the clinical history.

After arraying scans for each individual chronologically, each clinical event was scored by a doctor as primary acute, possible, follow-up, chronic, no valid result, secondary or negative (see supplementary material). A new clinical event was defined by a new illness leading to a hospital visit. The date of onset of the event was assigned as the date of onset of symptoms where this was recorded in the scan report, otherwise as the earliest date of hospital admission or scan. The reviewing doctor had no access to vaccination status except where a mention of this was embedded in the scan report. A second doctor re-scored all scans that had been coded non-negative and a random sample of negative scans, with the scoring of the first reviewer masked.

### Ascertainment of CVT from discharge diagnoses and death certificates

Records of all hospital discharges in Scottish Morbidity Record 01 (SMR01) from 1 December 2020 to 2021-04-28 and National Register of Scotland death registrations from 1 December 2020 to 2021-04-16 were queried for any mention of a CVT code as discharge diagnosis or cause of death. The ICD-10 codes used were: I63.6 (cerebral infarction due to cerebral venous thrombosis, nonpyogenic); I67.6 (nonpyogenic thrombosis of intracranial venous system); and G08 (intracranial and intraspinal phlebitis and thrombophlebitis). Records with mention of a local secondary cause such as infection, intracranial abscess or brain tumour were excluded by matching of the regular expression `^A|^B|^C7[012]|^D3[23]|G06[02]` in any of the diagnostic codes.

### Statistical methods

The case-crossover design compares event rates in time windows of recent and less recent exposure; this is equivalent to a matched case-control study in which the case and the control are the same person in different time windows [12]. We pre-specified that the time window for recent exposure was to be defined as 1 to 14 days after vaccination. For comparison with other studies that have used a 28-day time window, we also tabulated events and rates for the period 15-28 days. All post-vaccination cases were within 84 days of vaccination, so for the case-crossover analysis the 1-14 day time window is compared with the 15-84 day time window. For each vaccine product and each time window, the number of person-fortnights at risk was calculated to the end of follow up. To allow for the variation between health boards in latest date of case ascertainment via scans, the person-days in each exposure category from 14 April to 17 May 2021 were multiplied by 0.917 (the proportion of the population not covered by Forth Valley or Dumfries & Galloway where only records up to 14 April were extracted) and the person-days from 17 May 2021 to 1 June 2021 were multiplied by 0.121 (the proportion covered by Lanarkshire where records up to 1 June 2021 were extracted).

As the numbers of observed events were too small for confidence intervals to be equivalent to credible intervals, Bayesian credible intervals were calculated directly from the quantiles of the posterior distribution. With a flat prior on the logarithm of the event rate and *r* observed events in *n* million person-fortnights, the posterior distribution of the event rate is a gamma distribution with shape parameter *r* and inverse scale parameter *n*. Conditional on the total number of events, the likelihood given *r*_1_ events in *n*_1_ person-days in the recently exposed time window and *r*_0_ events in *n*_0_ person-days in the less recently exposed time window is 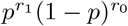 where *p* = *n*_1_*e*^*β*^*/*(*n*_0_ + *n*_1_*e*^*β*^) and *β* is the log rate ratio. With a flat prior on the log rate ratio *β*, normalizing this likelihood as a function of *β* gives the posterior distribution of *β*.

## Results

### CVT case ascertainment

As shown in Supplementary Figure S1, 142964 neuroimaging reports pertaining to 80905 individuals were retrieved. After de-duplication and retention of all venogram reports, and any reports containing keywords as described in the Methods section, 2760 scans pertaining to 1607 individuals were retained for manual coding. Table 1 shows the codes assigned to these individuals after review. The independent coding validation by a second masked coder resulted in only one reassignment where a case with metastatic cancer but no brain metastases was reassigned as primary acute rather than secondary. 48 cases ascertained through scans were coded as having a primary acute CVT after 1 December 2020, and 18 were coded as “possible CVT”. No additional cases in the study period were ascertained through death certificates with CVT codes.

**Table 1.** Scoring of CVST diagnostic categories on 1605 individuals with scans after 1 December 2020 that were filtered by study description or text strings in report for manual review

|  | Number of individuals |
| --- | --- |
| Primary acute | 48 |
| Possible | 18 |
| Chronic | 19 |
| Follow-up | 10 |
| No valid result | 0 |
| Secondary | 33 |
| Negative | 1479 |
| All | 1607 |

The yield of cases through SMR01 discharge reports up to the latest discharge in the SMR01 extract is shown in Table 2 In the extract used for this study there were 34 individuals with first mention after 1 December 2020 of a CVT code, of whom 29 had no mention of a secondary code on any discharge record. Of these 29, 7 had neuroimaging reports that showed a secondary cause, chronic thrombus, or stenosis rather than thrombus. Only 3 of the 29 events ascertained from SMR01 discharge records had no neuroimaging report retrieved by the PACS query. Of these one had an unrelated condition as main diagnosis, one had a CVT code as main diagnosis but had stayed only one night in hospital, and one had a CVT code as main diagnosis and a scan record but no report, from one of the two health boards that did not include the text of scan reports in the PACS database. On the basis that CVT diagnostic code as main condition had high specificity (14/17) the two cases with CVT coded as main condition were retained as primary acute. As none of these cases without scan reports were post-vaccination, their inclusion or exclusion does not affect the case-crossover results.

**Table 2.** 29 SMR01 discharges with CVT diagnoses from 1 December 2020 by scan report

|  | Primary acute | Chronic / secondary / negative | No scan | All |
| --- | --- | --- | --- | --- |
| As main condition | 14 | 3 | 2 | 19 |
| As other condition | 5 | 4 | 1 | 10 |
| Total | 19 | 7 | 3 | 29 |

Ascertainment based on mention of CVT codes G08, I63.6, I67.6 excluding records with mention of codes for secondary causes as described in Methods

Table 3 shows that of the 48 primary acute CVT cases ascertained through neuroimaging, 19 had a discharge record with a CVT code, 8 had a discharge record with no CVT code, and 21 had no discharge record. Note that as coding of hospital admissions is done after discharge with varying lags, no exact cutoff date for the latest date of admission of cases ascertained through SMR01 can be defined. However as shown in 3 of 35 primary acute CVT case ascertained through PACS up to the end of the first quarter of 2021 (a period probably fully captured by the SMR01 extraction), only 17 had had a CVT diagnosis code on an SMR01 record.

**Table 3.** 48 scans coded as primary acute by 3-month calendar period and mention of a CVT code on discharge record

|  | CVT diagnosis | Other diagnosis | No coded discharge record | All |
| --- | --- | --- | --- | --- |
| 2020-Q4 | 8 | 1 | 2 | 11 |
| 2021-Q1 | 9 | 7 | 8 | 24 |
| 2021-Q2 | 2 | 0 | 11 | 13 |
| Total | 19 | 8 | 21 | 48 |

Combining the 49 cases ascertained through scans and scored as primary acute with the two additional cases with CVT coded as main diagnosis that had been ascertained only through SMR01, 50 cases classified as primary acute were retained for analysis, together with 18 scan-ascertained cases coded as possible.

### Relation of CVT to vaccine exposure

Table 4 shows the number of doses by age group and vaccine product from 1 Dec 2020 to 2021-07-15. The vaccine product was missing for about 0.4% of recorded doses.

**Table 4.** Vaccine doses by age group up to 2021-07-15

|  | Pfizer | AstraZeneca | Moderna | Total doses |
| --- | --- | --- | --- | --- |
| 0-39 | 1035856 (39%) | 378507 (9%) | 144157 (84%) | 1558520 (23%) |
| 40-59 | 690208 (26%) | 1837077 (46%) | 27208 (16%) | 2554493 (37%) |
| 60 or more | 963420 (36%) | 1815596 (45%) | 862 (1%) | 2779878 (40%) |
| . | 4 (0%) | 1113 (0%) | 0 (0%) | 1117 (0%) |
| Total doses | 2689488 | 4032293 | 172227 | 6894008 |

Table 5 tabulates the post-vaccination cases by vaccine product and time since vaccination. As pre-specified, the time window of 1 to 14 days was used to define recent exposure. All other events in vaccinated individuals occurred between 15 and 84 days after vaccination. Case-crossover estimates of rate ratio were based on comparing the 1-14 day time window with the 15-84 day time window. In each time window, the number of person-fortnights exposed in that time window is the denominator from which the rate is calculated.

**Table 5.** CVT cases after vaccination, by time since vaccine dose

| Product | Days since vaccination |  |  | Rate ratio |
| --- | --- | --- | --- | --- |
|  | 1-14 | 15-28 | 29-84 | 1-14 vs 15-84 |
| **Person-fortnights exposed** |  |  |  |  |
| Pfizer | 1908751 | 1784505 | 4942003 | . |
| AstraZeneca | 3154182 | 2698048 | 7390991 | . |
| Moderna | 23578 | 17801 | 21013 | . |
| **Cases scored as primary acute** |  |  |  |  |
| Pfizer | 2 | 1 | 0 | . |
| AstraZeneca | 7 | 2 | 5 | . |
| Moderna | 0 | 0 | 0 | . |
| Rate (95% credible interval) primary acute per million person-fortnights |  |  |  |  |
| Pfizer | 1 (0.1 to 2.9) | 0.6 | 0 | 7 (0.6 to 95.8) |
| AstraZeneca | 2.2 (0.9 to 4.1) | 0.7 | 0.7 | 3.2 (1.1 to 9.5) |
| Moderna | 0 . | 0 | 0 | . |
| **Cases scored as primary acute or possible** |  |  |  |  |
| Pfizer | 2 | 3 | 1 | . |
| AstraZeneca | 9 | 3 | 7 | . |
| Moderna | 0 | 0 | 0 | . |
| Rate (95% credible interval) primary acute or possible per million person-fortnights |  |  |  |  |
| Pfizer | 1 (0.1 to 2.9) | 1.7 | 0.2 | 1.8 (0.2 to 8.9) |
| AstraZeneca | 2.9 (1.3 to 5) | 1.1 | 0.9 | 2.9 (1.1 to 7.2) |
| Moderna | 0 . | 0 | 0 | . |

17 definite CVT events and 25 definite or possible CVT events occurred post vaccination: the others were in individuals who had not been vaccinated at the time of onset. Of the 7 cases with onset within 14 days of exposure to AstraZeneca vaccine, 6 were in people aged less than 60 years and 4 were in women.

The observed rate of definite (primary acute) CVT diagnoses in the 14 days after exposure to AstraZeneca vaccine is estimated as 2.2 (95% credible interval 0.9 to 4.1) per million doses. For the Pfizer vaccine, the corresponding rate was 1 (95% credible interval 0.1 to 2.9) per million doses. There were no cases in the small numbers exposed to Moderna vaccine. For comparison, the observed rate of primary acute CVT in the 15-84 day time window after exposure was 12 per million per year (95% credible interval (5.3 to 22.2). The rate ratio associated with exposure in the last 14 days to the AstraZeneca vaccine was estimated as 3.2 (95% credible interval 1.1 to 9.5) for primary acute CVT, and 2.9 (95% credible interval 1.1 to 7.2) for primary acute or possible cases.

For comparison with other studies that used a 28-day time window, the rate of primary acute CVT within 28 days of vaccination with the AstraZeneca product, based on 9 events in 2926114 28-day time intervals, was 3.1 (95% credible interval (1.4 to 5.4) per million doses. In those aged under 60 years the rate, based on 8 events in 1319795 28-day time intervals was 7 (95% credible interval (3.1 to 11.9) per million doses.

### Checking for bias in case ascertainment

Fig 1 shows the total number of venograms (panel a) and other head scans (panel b) in boards with complete ascertainment using PACS, by 7-day sliding window of study date. The average number of venogram studies was about 50 /day from December 2020 to March 2021, but increased sharply in the first two weeks of April to a peak of nearly 150 /day day in mid-April, around the time that the association of CVT with vaccination first received wide publicity in the UK. However the number of individuals exposed to the AstraZeneca vaccine in the last 14 days had peaked earlier in late March. Of the 9 cases that occurred within 28 days of vaccination with the AstraZeneca product, 6 were before April 2021, when the increase in number of venograms per day began.

**Fig 1.**
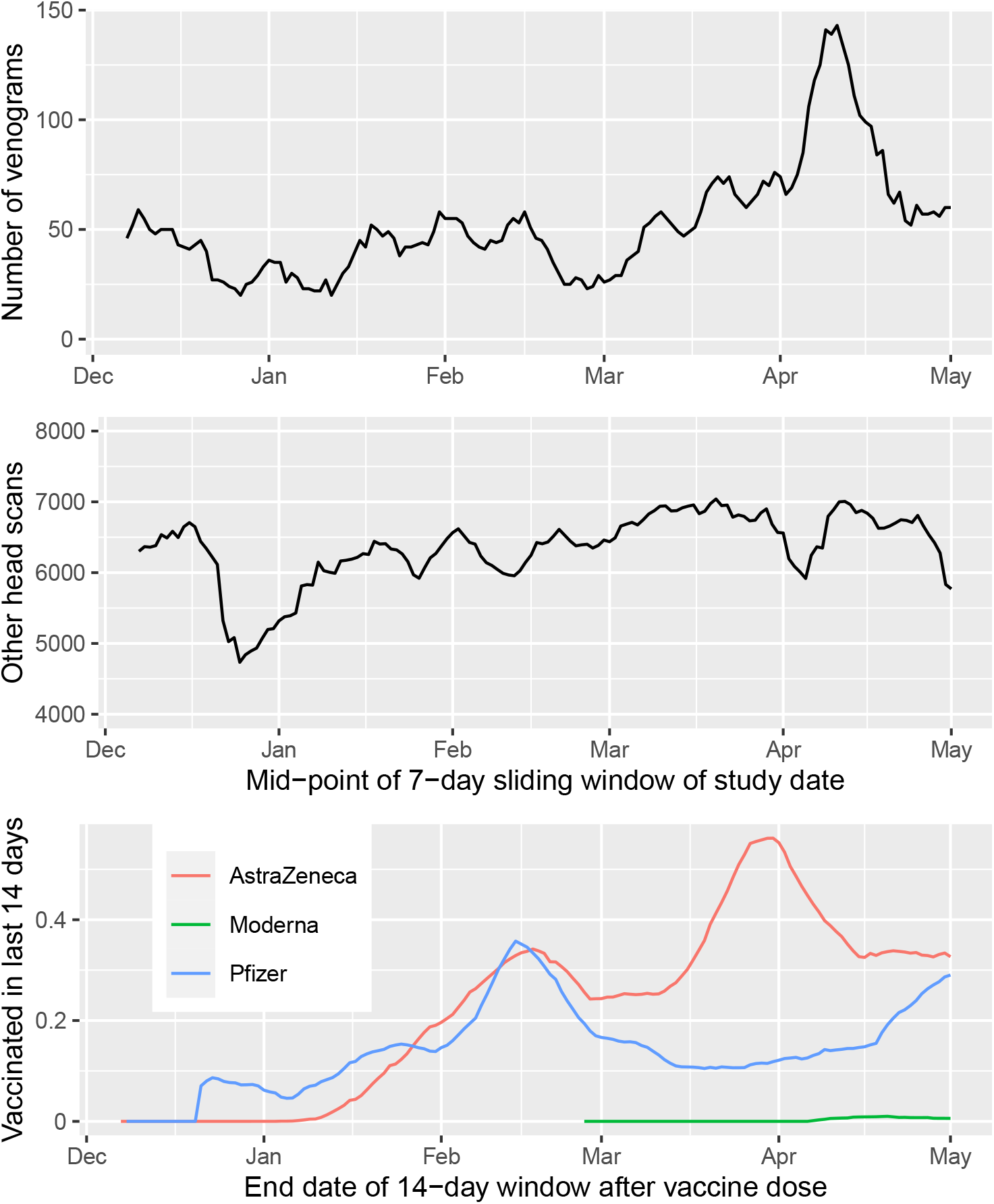
(a) Venograms in the 11 PACS boards by 7-day sliding window of dates; (b) Other head scans; (c) Population recently exposed: millions vaccinated in last 14 days

## Discussion

### Statement of principal findings

In this population-wide study we have captured all diagnosed CVTs nationally since before the start of the vaccination programme. We have shown that ascertainment of CVTs from diagnostic codes on hospital discharge records has low sensitivity for CVT; systematic review of scan reports is required for complete ascertainment of diagnosed cases and to distinguish primary acute from secondary cases. The number of events in those recently exposed to vaccine is small but we can still calculate a credible interval for the incidence rate and thus an upper bound. For the first 14 days after AZ vaccination, we calculate an upper bound of 4.9 per million doses on the absolute rate, and 9.1 for the rate ratio. Although the rate of venogram studies increased markedly in early April 2021 after the association of CVT with vaccination had received wide publicity, most of the vaccine-associated cases in this study were diagnosed before this, and thus it is unlikely that ascertainment could have biased the association with recent vaccine exposure. When combined with prior evidence of excess risk of CVT associated with the AstraZeneca vaccine in other populations, the case-crossover analysis supports a causal interpretation of this association. Although the number of cases exposed to the Pfizer vaccine is too small for any comparison between the two products, an upper bound can still be placed on the absolute risk associated with the Pfizer vaccine.

### Strengths and limitations of this study

A strength of this study is the complete ascertainment of cases in the population, not reliant on adverse event reporting or diagnostic coding of hospital discharge records. Manual review of scan reports, which typically included a summary of the clinical history allowed some CVT cases to be recoded as secondary to a local lesion such as a tumour, allowed some to be recoded as chronic rather than acute CVT, and allowed the date of onset to be determined accurately. Accurate assignment of date of onset is required for the case-crossover analysis to be valid.

The case-crossover design eliminates confounding by time-invariant factors, which may be strong where vaccine allocation is based on pre-existing risk conditions that were used to allocate priority for vaccination. This has allowed us to show that for the Astra Zeneca there is evidence of a causal association of CVT with vaccine exposure. Note that our analysis assumes an ignorance prior; it ignores any prior evidence of association.

Limitations of our study are that we do not have access to data on platelet counts, D-dimer or platelet factor 4 antibody levels, allowing enumeration of the number of CVT events that are part of the formally defined VITT / TTS syndrome as meeting associated haematological criteria. We note that the Brighton Collaborative however acknowledges that the TTS syndrome definition may be too restrictive by excluding isolated thrombotic events that are causally related to vaccine. Also we did not ascertain venous thromboses at sites other than brain.

### Relation to other studies

From this study the background incidence of primary acute CVT in Scotland (excluding the 14-day time window after vaccination) can be estimated as about 12 per million adults per year: this is similar to the estimate of 16 per million per year in Australia in 2016 [13] based on ascertainment of cases via neuroimaging records. Most other estimates of CVT incidence have relied on ascertainment via diagnostic coding in health informatics systems, which is likely to underestimate the incidence of CVT. A recent study from Scotland based on primary care, hospitalisation and death records in the total population reported 19 CVT events between 8 December 2020 and 14 April 2021 [8]. For the same period our study ascertained 41 primary acute cases of CVT in Scotland.

Adverse event reporting systems can give early warning of unexpected effects but cannot be relied on to estimate incidence rates. By early April 169 CVTs had been reported to the EMA by which time about 34 million doses of Astra Zeneca vaccine had been administered, giving a rate of about 5 per million doses. In the UK as of 9 June 2021 the Yellow Card scheme operated by the MHRA had received 390 reports of cases of “major thromboembolic events with concurrent thrombocytopenia”. Cerebral venous sinus thrombosis was reported in 140 of these. By 2 June 2021 30.1 million doses had been administered, so allowing for a one-week lag the incidence was 13.0 per million doses for the syndrome and 4.7 per million doses for CVT [14], Our estimate of 3.5 per million doses in the first 28 days is similar to that estimated by the MHRA, indicating that the Yellow Card system, for all its limitations, has not seriously underestimated the incidence of vaccine-associated CVT.

The upper bound of 6.2 per million doses for incidence of CVT within 28 days of vaccination with the AstraZeneca product estimated in this study is rather lower than estimates reported from Scandinavia and Germany. In a study of all 281264 individuals aged 18-65 years in Denmark and Norway who received a first dose of the AstraZeneca vaccine, there were were 7 observed CVT events within 28 days of vaccination compared with 0.3 expected from rates in the general population [7]: a rate of 25 per million doses. A study of CVT events ascertained through neurologists from nine states in Germany where 2320535 first doses had been administered reported 27 cases within 31 days of AstraZeneca vaccine recipients, giving a rate of 15 per million doses within 31 days of vaccination [15]. In those aged under 60 years the rate in the German study was 18 per million doses, compared with 7 per million doses for the same age group and 28-day time interval. The reasons for this difference between the UK and EU countries are not clear. A report from the European Medicines Agency dated 24 March 2021 stated that they had requested AstraZeneca to provide “a full batch analysis for specific lots and batch data from UK supplied lots to understand if there are any clear differences between that and the EU products” [16].

### Policy implications

This study based on complete ascertainment of CVT cases makes it possible to set a definitive upper bound on the rate of vaccine-associated CVT in Scotland. The results reinforce the importance of establishing comprehensive surveillance of adverse events occurring after vaccination. By using e-health record systems we were able to obtain all neuroimaging reports for the population and to report preliminary results to public health agencies within a few weeks of initiating this study. This entailed labour-intensive manual coding of scan reports. For longer-term surveillance and scaling to larger populations a natural language processing algorithm could be developed to identify CVTs in imaging reports, but some manual coding would still be required especially to assign the date of onset.

Policy on the continued use of the AstraZeneca vaccine has been driven by estimates of the risk/benefit ratio, with risk of TTS estimated from adverse event reporting schemes [17]. Thus on 7 May 2021 the Joint Committee on Vaccination and Immunisation advised that those under age 40 should be offered an alternative to the AstraZeneca vaccine [18]. Evaluating the risk / benefit ratio of COVID-19 vaccination in healthy young adults and children depends on being able to detect rare adverse events post-vaccination through surveillance, so that the risk of such events can be compared with the low risk of severe complications of COVID-19 in these groups.

## Data Availability

The component datasets used here are available via the Public Benefits Privacy Panel for Health at https://www.informationgovernance.scot.nhs.uk/pbpphsc/ for researchers who meet the criteria for access to confidential data. All source code used for derivation of variables, statistical analysis and generation of this manuscript is available on https://github.com/pmckeigue/covid-scotland_public

## Declarations

### Public and Patient Involvement statement

This study was conducted under approvals from the Public Benefit and Privacy Panel for Health and Social Care which includes public and patient representatives.

### Ethics approval and information governance

This study was performed within Public Health Scotland as part of its statutory duty to monitor and investigate public health problems. Under the UK Policy Framework for Health and Social Care Research set out by the NHS Health Research Authority, this does not fall within the definition of research and ethical review is not required. Individual consent is not required for Public Health Scotland staff to process personal data to perform specific tasks in the public interest that fall within its statutory role. The statutory basis for this is set out in Public Health Scotland’s privacy notice.

A Data Protection Impact Assessment (DPIA) allows Public Health Scotland staff to link existing datasets. This study was approved under COVID-19 Rapid DPIA 20210023.

### Transparency declaration

HC as the manuscript’s guarantor affirms: that the manuscript is an honest, accurate, and transparent account of the study being reported; that no important aspects of the study have been omitted; and that any discrepancies from the study as originally planned and registered have been explained. This manuscript has been generated directly from the source data by a reproducible research pipeline.

### Contributors

PM, RB, DM and HC provided substantial contributions to the conception or design and drafting of the manuscript. JB provided substantial contributions to the data acquisition. CR, JM and MO provided substantial contributions to the interpretation. All authors contributed to revising the manuscript critically for important intellectual content and approved the final manuscript.

## Acknowledgements

We thank Matteo Abideni (Philips Ltd), Alan Fleming (National Services Scotland), and Kenny Scott (National Services Scotland) for help with querying the PACS database

## Supplementary Material

**Fig S1.**
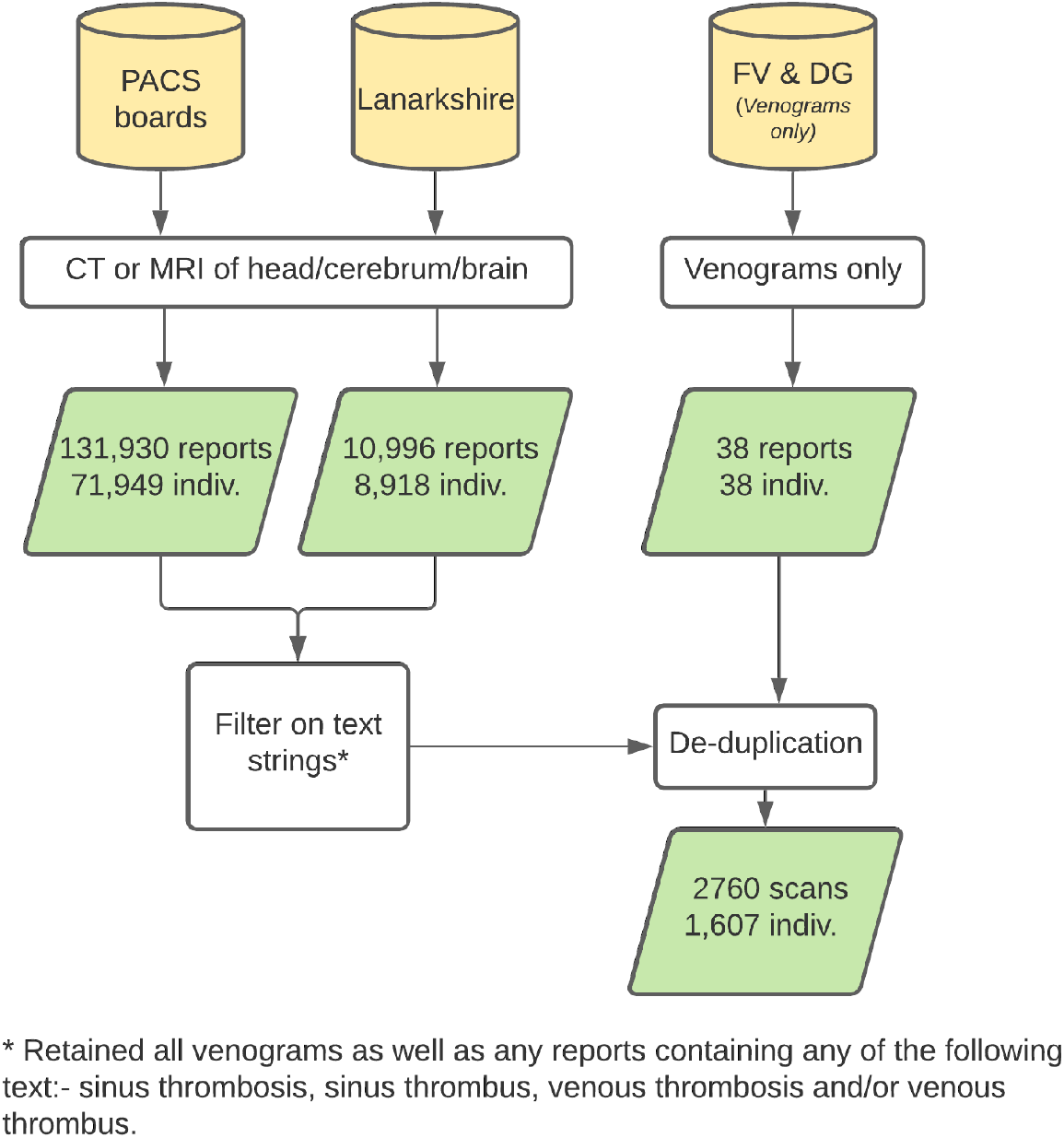
Flow diagram for case ascertainment from scan reports

## PACS query for ascertainment of scans possibly informative for CVT

○ (Modality=CT OR Modality=MR) AND (Study description contains “head” or “cerebr*” or “brain”) **OR**
  ○ RIS code = ANY OF

- CVENO CT Venogram
- CVEIC CT Venogram Intracranial
- CVECE CT Venogram cerebral
- CTHVG CT Head Venogram
- MVENO MRV Venogram
- CBNTA CT Brain neck thorax Abdo and pelvis
- CHAP CT Head abdomen and pelvis
- CHAPC CT Head abdomen pelvis with contrast
- CHNTAP CT Head neck thorax abdomen and pelvis
- CHTA CT Head thorax and abdomen
- CHTAP CT Head thorax abdomen and pelvis
- CHTAPC CT Head thorax Abdo pelvis with contrast
- CHTH CT Head and thorax
- CHTHAC CT Head thorax abdomen with contrast
- CHTHC CT Head and thorax with contast
- CSKNE CT Head and neck
- CSKPE CT Brain perfusion study
- CSKUH CT Head
- CSKUHC CT Head with contrast
- CSKUC CT Head with contrast
- MAICA MRA Head
- MAICAC MRA Head with contrast
- MBRCC MRI Brain and cervical cord
- MSKPE MRI Head brain perfusion study
- MSKUH MRI Head
- MSKUHC MRI Head with contrast
- MSKUS MRI Head spectroscopy
- MVSKU MRV Cerebral veins

## Protocol for scoring neuroimaging studies

### Scoring at scan level

- **negative** - no evidence of any venous sinus thrombosis in the report
- **secondary** - evidence of acute or chronic CVT but there is another local pathology contributing such as meningioma, mastoiditis, sinusitis or encephalitis. Systemic factors such as Factor V Leiden deficiency, post-partum state, use of oral contraceptives, or pro-thrombotic drugs are not classified as secondary causes. Trauma caused by a external event such as traffic accident or assault is classified as a secondary cause, but history of a fall that could have resulted from a primary brain event is not classified as a secondary cause.
- **chronic** - this code is assigned when there is a new presentation but the scan shows evidence of unchanged or resolving thrombosis or recanalising thrombosis. There may or may not be a definitive history of an earlier acute primary event.
- **follow-up** - this code is assigned where the scan shows evidence of thrombosis but has been done as a follow up of a prior event either during the same hospitalisation or later as a routine follow-up. This code was assigned to reports without the primary scan necessarily being available at time of report coding.
- **possible** - this code is used where there is some evidence that may suggest thrombosis but which is not definitive; many such reports will recommend further investigation
- **primary acute** - scan reported as consistent with venous thrombosis, not assigned as chronic and which cannot definitely be assigned to other local cause.
- **no valid result** - there is a report but the report text is missing or the scan was declared to be a technical failure or unreadable

### Coding at person level

For this process all available scans extracted for an individual were arrayed at person level. An event encompasses all scans pertaining to a new presentation. A follow up scan done several months or years later is a new event.

- **negative** - none of the scans have found evidence of a CVT or an initial scan with possible CVT is followed by a definitive scan such as venogram that rules out CVT
- **secondary** - an event where there is evidence of acute or chronic CVT but there is another local brain pathology contributing such as meningioma or mastoiditis or sinusitis or encephalitis
- **chronic** - there is a new presentation and one of the scans shows chronic thrombus
- **follow-up** - Most events that include follow up scans during that admission will be coded as primary acute events with the date of the event being that of the presentation date of the originating thrombus event. For follow up scans in a separate hospital attendance these are coded as follow up but the event onset date is given as the original primary event date where it is available. Where the primary event is not referred to or given a date the final event code maybe left as follow-up with the date of onset left blank
- **possible** - one of the scans for the event found evidence of a possible CVT and no subsequent scan ruled this out or resolved whether or not a thrombus was present
- **primary acute** – a new presentation where any scan shows changes consistent with thrombus that is not assigned as chronic and is not attributed to a secondary local cause

### Examples of how the protocol is applied

1. Patient presents with a seizure – all scans are negative. Two weeks later they present with new onset confusion – CT scan negative, MR venogram shows bleed and CVT. The first admission is coded as negative for each scan and for the event. The second set of scans is coded as negative and primary acute respectively and the event is coded as primary acute.
2. Patient presents with confusion and headache after a fall – CT head has no abnormality, subsequent CT venogram shows filling defect in venous sinus – a scan three days later shows similar picture. The reports are coded as negative, primary acute, follow up. The event is coded as primary acute.

